# Alcohol, calories and obesity: A rapid systematic review and meta-analysis of consumer knowledge, support and behavioural effects of energy labelling on alcoholic drinks

**DOI:** 10.1101/2020.08.25.20181479

**Authors:** Eric Robinson, Gabrielle Humphreys, Andrew Jones

## Abstract

Mandatory energy (calorie) labelling of alcoholic drinks is a public health measure that could be used to address both alcohol consumption and obesity. We systematically reviewed studies examining consumer knowledge of the energy content of alcoholic drinks, public support for energy labelling and the effect of energy labelling of alcoholic drinks on consumption behaviour. Eighteen eligible studies (from 16 sources) were included. Among studies examining consumer knowledge of the energy content of alcoholic drinks (N=8) and support for energy labelling (N=9), there was moderate evidence that people tend to be unaware of the energy content of alcoholic drinks (pooled estimate: 74% [95% CIs 64-82%] of participants inaccurate estimating energy content) and support energy labelling (pooled estimate: 64% [95% CIs 53%-73% support policy]. Six studies examined the effect of energy labelling on consumer behaviour and findings were indicative of no likely effect of labelling. However, the majority of studies were of low methodological quality, used proxy outcome measures and none of the studies were conducted in real-world settings, resulting in a very low level of evidence. Further research is required to determine whether energy labelling of alcoholic drinks affects consumer behaviour and is likely to be an effective public health policy.

## Introduction

Excessive alcohol consumption produces a large global burden on health ^1^ and has been found to be consistently associated with increased risk of developing a range of health conditions, including liver and cardiovascular disease ^2 3^. Although findings to date have been mixed, a number of studies suggest that heavier alcohol consumption may be a risk factor for weight gain and obesity ^4^. Amongst regular drinkers, energy derived from alcohol can make a significant contribution to daily energy intake ^5^. For example, a Canadian study of regular alcohol drinkers found that the average participant consumed 250 calories from alcoholic drinks per day and this accounted for 11% of their daily energy requirements ^6^. Furthermore, laboratory evidence suggests that individuals do not compensate in the short term for the calories in alcohol by consuming less food ^7^. There is also some research which has concluded that people are unaware of the number of calories in alcoholic drinks ^8^, which may suggest that provision of energy information on alcoholic drinks may help consumers to reduce their alcohol consumption and daily energy intake.

At present, in the UK and EU, manufacturers do not have to provide nutritional information, such as number of calories per serving, on alcoholic drinks by law ^9 10^. Research also indicates that previous voluntary pledges made by the alcohol industry to provide nutritional information on alcoholic beverages have been ineffective ^11^. There have been calls made by public health bodies to make labelling of calorie information on alcoholic drinks mandatory ^12^ and in 2020, the UK government announced an intention to consult on a mandatory calorie labelling of alcoholic drinks policy as part of their public health strategy to reduce obesity ^10^. The effectiveness of a mandatory calorie labelling policy will be in part determined by whether calorie information is likely to be informative for consumers (e.g. do consumers already know how many calories are in drinks?), public acceptability of the policy ^13^ and the effect that labelling has on consumer behaviour ^14^. A number of systematic reviews have examined the effect that nutritional labelling of food products has on dietary behaviour ^15 16^. However, there has been no systematic review of research on energy labelling of alcoholic drinks and it is unclear the extent to which consumers are aware of the number of calories in alcoholic beverages, whether mandatory energy labelling of alcoholic drinks is acceptable (policy support) and whether energy labelling of alcoholic drinks affects consumer behaviour (e.g. reduces energy intake). Therefore, the aim of the present research was systematically review existing evidence on consumer knowledge of the energy content of alcoholic drinks, consumer support for energy labelling of alcoholic drinks and consumer effects of energy labelling of alcoholic drinks.

## Method

### Rapid review approach

As we conducted this research in response to an announcement of a consultation and call for scientific evidence on a mandatory calorie labelling policy by UK government, we used rapid systematic review methodology ^17^. Rapid reviews are used to provide timely and relevant evidence synthesises to inform policy and practice whilst maintaining the rigour and reproducibility of traditional systematic review methodology ^18^. Rapid reviews typically achieve this by using expedited review processes ^18 19^, such as limiting eligibility of research to published articles only, searching a limited number of electronic databases or by reducing the number of researchers conducting the review (e.g. cross-checking of a proportion of extraction as opposed to independent extraction by a second author).

### Eligibility criteria and study selection

We included studies that had examined consumer knowledge of the energy content of alcoholic drinks, consumer support for energy labelling of alcoholic drinks and/or the effects of alcoholic drink energy labelling on consumption behaviour. Published journal articles were eligible for inclusion. Research reports published by public health bodies/research agencies that had not been published in an academic journal, but included a sufficient level of detail to allow eligibility to be assessed (i.e. study methodology and results section included in report) were also eligible. Due to the scope of the project we did not include unpublished papers (e.g. student dissertations). Due to the study outcome measures that were eligible (see below), studies that included only qualitative analyses were not be eligible for inclusion. However, if a study used a qualitative approach but also included quantitative data relating to an eligible outcome (e.g. % of participants accurately identifying number of calories in an alcoholic drink), the study was eligible for inclusion. The pre-registered protocol for the review is available at https://osf.io/8gpm5/ and is registered on PROSPERO (CRD42020203817).

#### Populations

No exclusion criteria on study settings or sampling method used to recruit participants were used, with the exception of excluding studies that had sampled participant groups on the basis of their professional status. For example, studies on consumer support for energy labelling of alcohol drinks that had sampled alcohol industry employees may not be representative of the general public.

#### Interest/Intervention

For studies examining consumer knowledge of the energy content of alcoholic drinks, at least one self-report measure of perceived energy content (e.g. perceived/estimated calorie content of an alcoholic drink) was required. For studies examining consumer support for energy labelling, studies were required to have included at least one self-report measure relating to policy support (e.g. ‘Do you think there should be calorie labelling of alcoholic drinks?’’). For studies that examined consumer effects of energy labelling of alcoholic drinks, studies were required to have examined the effect of providing energy information about alcohol drinks (e.g. calorie information added to labels) on a measure relating to alcohol or food consumption behaviour. For completeness, we also included studies that examined the effect of alcohol energy labelling on consumer knowledge of the energy content of alcohol drinks and/or support for energy labelling of alcoholic drinks.

#### Comparison

For studies examining the consumer effects of energy labelling of alcoholic drinks, studies were required to include one group that provided participants with energy labelling (with or without additional nutritional information) and a comparator group that had not received energy/nutrition labelling.

#### Outcomes

For studies examining knowledge of the energy content of alcoholic drinks, eligible outcome variables were directional accuracy scores (i.e. average (bi)directional difference between estimated calorie content and actual content) and/or % of participants accurate (i.e. % of sampled participants that estimated the ‘correct’ number of calories in an alcoholic drink). Measures that did not allow for a quantifiable measurement of accuracy were ineligible (e.g. extent to which participants agree whether a drink has a large number of calories). For studies that examined consumer support for energy labelling, eligible outcome variables were % of participants endorsing support for the policy (i.e. % selecting ‘yes’ or ‘agree’) or average level of support (i.e. score on a 1-7 scale, from no support to complete support). Measures relating to perceived efficacy of energy labelling (e.g. ‘I think most people would drink less if energy labelling was on alcoholic drinks’) are not a direct measure of policy support and were therefore ineligible. For studies examining consumer effects of energy labelling of alcoholic drinks, eligible outcome variables were objective or self-report measured alcohol or food consumption behaviour (e.g. amount of alcohol consumed) and related ‘proxy’ behavioural outcomes (e.g. self-reported consumption or purchasing intentions or hypothetical choice). Measures that were not directly related to consumption (e.g. self-reported liking or sensory evaluations of beverage) were ineligible.

### Article identification strategy

We searched PUBMED and Scopus (no date limits) for published articles in peer reviewed journals. To maximise coverage of all likely relevant literature we used the following search terms: (‘Nutrition’ OR ‘Calorie’ AND ‘Labelling’) AND (‘Alcohol’ OR ‘Ethanol’ OR ‘Beer’ OR ‘Cider’ OR ‘Wine’ OR ‘Spirits’ OR ‘Cocktails’). One author conducted the initial title and abstract screening to exclude articles clearly unrelated to the review aims. Two independent authors conducted the full-text screening to determine final eligibility. For all eligible articles identified through searches, one author used forward citation tracking (Google Scholar) and searched reference lists to identify any further articles. A second author verified eligibility of articles identified through citation tracking and reference list searching.

### Data extraction

For each study one author extracted the following information, and a second checked all extraction for accuracy: Bibliographic information, Information on country; Participant group sampled (e.g. university students, online panel, local community) and data collection setting (e.g. laboratory study), Summary information on participant age, gender, education level, alcohol drinking habits and body mass index (BMI), Whether the study examined i) consumer knowledge of the energy content of alcoholic drinks, ii) consumer support for energy labelling of alcoholic drinks and/or iii) consumer effects of energy labelling of alcoholic drinks, eligible outcome variables used, Results relating to eligible outcome variables of interest, including descriptive statistics and results of any relevant statistical analyses. For experimental studies that assigned participants to view vs. not view information about the energy content of alcoholic drinks: details of the information that participants were exposed to in each condition and procedural information on how information was presented.

### Study quality indicators

Because eligible studies were expected to vary in design and address different research questions, we developed a review specific checklist to assess study quality and risk of bias based on existing study quality checklists and criteria ^20-23^. The following 10 study quality indicators were assessed: 1) Was the study sample size justified and was this justification adequate*?* 2) Was the study sample size very small (< 20 participant per group for an experimental study, < 30 participants for observational study)? 3) Was the study methodology described in sufficient detail? 4) Were study results described in sufficient enough detail to support conclusions? 5) Were outcome measures appropriate for research question? 6) Was the study methodology and analysis plan pre-registered? 7) Where potential conflicts of interest reported? 8) For experimental studies examining consumer effects of energy labelling, were randomization methods used (and described) to allocate participants to conditions? 9) For experimental studies examining consumer effects of energy labelling, were efforts made to minimize participant awareness of study aims? 10) For experimental studies examining consumer effects of energy labelling, is information provided on: participant awareness of study aims? For each study, one author extracted the information and extraction was checked for accuracy by a second author. For a detailed description and examples for criteria see the Appendix.

### Study quality evaluation

Based on the above quality criteria we rated studies as being ‘Low’, ‘Moderate’ or ‘High’ in overall methodological quality. Low quality studies were those that scored poorly on a significant number of the quality criteria. In particular, poor reporting of study methodological information or results alongside sub-optimal scoring on other study quality criteria resulted in a score of ‘Low’, as these factors make it difficult to draw conclusions from a study with confidence. ‘Moderate’ quality studies were classed as studies lacking in a small number of the individual quality criteria that could cumulatively influence confidence in conclusions, but there were major concerns over any individual study quality that have been identified that are likely to invalidate conclusions (conclusions can be made with some confidence). ‘High’ quality studies were any that scored perfectly on each individual quality criteria or had relatively minor methodological limitations that would be unlikely to invalidate conclusions (e.g. do not report a sample size justification, but have a very large sample size). Two authors independently rated each study and initial agreement was high (94%). Disagreements were resolved through discussion with a third author.

### Synthesis of evidence

We planned to synthesise studies narratively and summarise current evidence for each study type separately. We graded the overall level of evidence for conclusions made from each study type using the GRADE approach ^24^. GRADE results in an overall grading of as: high, moderate, low or very low, based on considering the quality of included studies, consistency of findings, indirectness of evidence (e.g. reliance on studies using proxy measures of consumer behaviour) and imprecision (e.g. studies having relatively few participants and wide confidence intervals). After completing data extraction, we identified that there were a sufficient number of studies with similar methodology and reporting of results addressing consumer knowledge of energy content (% participants accurate) and policy support (% of participants in support of policy). We therefore meta-analysed studies with this information to provide a pooled estimate for each outcome using Generic Inverse Variance random effects models (see supplementary materials for full information). We did not meta-analyse studies examining consumer effects of energy labelling as study outcomes differed between studies.

## Results

### Study selection

Electronic searches of PUBMED and Scopus returned 853 articles. After removal of duplicates (N=60), 793 search records were title and abstract screened. After removal of articles unrelated to the research question, a total of 39 articles were identified for full-text screening. Of these articles, 10 were deemed eligible ^25-34^. See Figure 1 for exclusions. A further six eligible articles ^8 35-39^ were identified through forward tracking of citations, reference list searches and the authors’ knowledge. In total, 16 articles were included in the review and from these articles a total of 18 studies were deemed eligible for inclusion.

**Figure 1.**
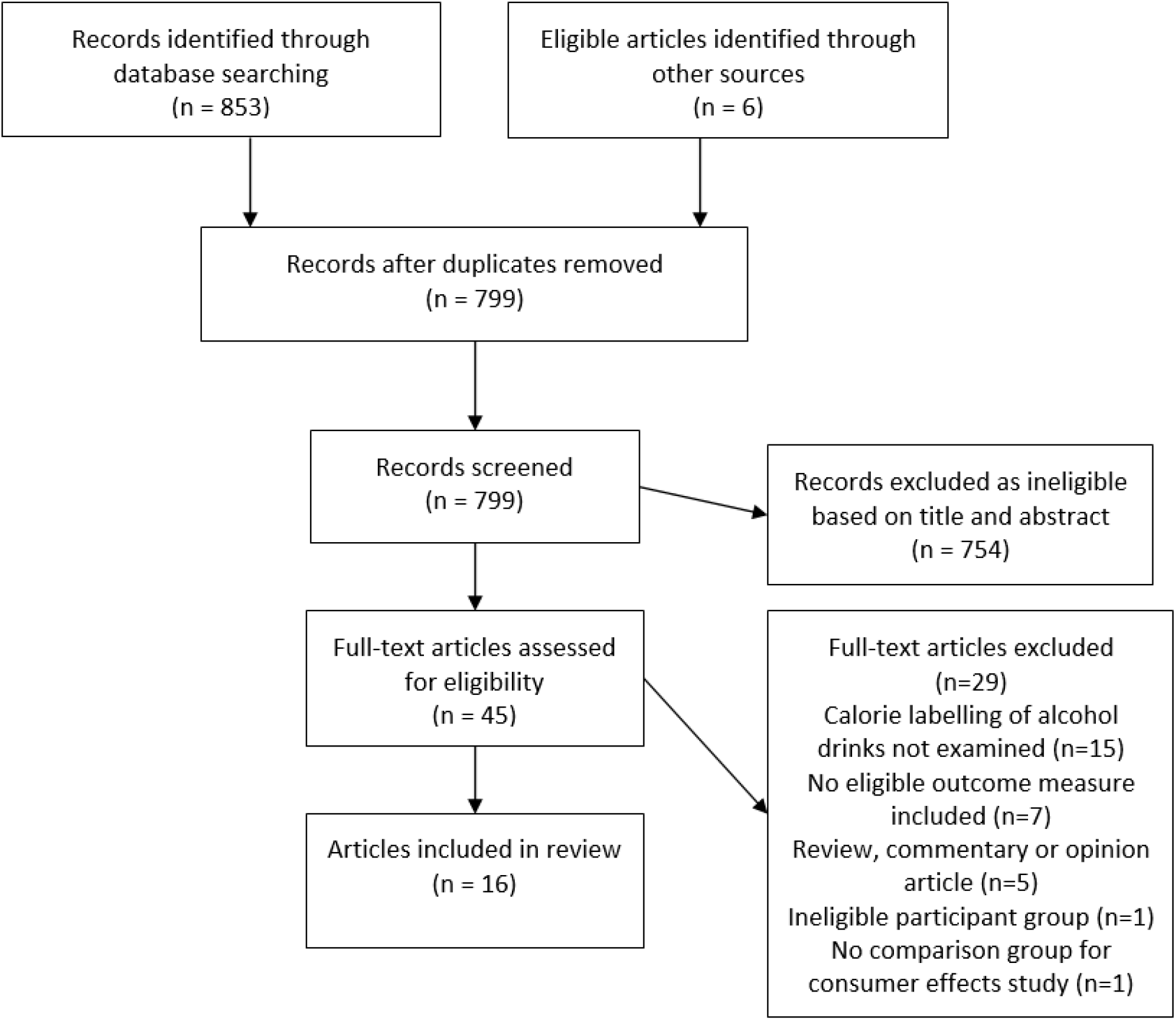
Study search and eligibility process.

### Overview of characteristics and quality of included studies

13 of the eligible articles were from published academic journals and 3 were reports from public health bodies/research agencies that were not published in academic journals (e.g. Alcohol Research UK). The majority of studies (n=18) included were of low methodological quality (n=13) and a minority were judged to be of moderate (n=4) and high (n=1) quality. Common reasons for low methodological quality ratings were insufficient procedural information, selective reporting of results, lack of conflict of interest information and no information concerning participant awareness of study aims in studies examining consumer effects of energy labelling. See Table A1 in the appendix for ratings of individual study quality criteria.

### Knowledge of energy content of alcoholic drinks

Eight studies examined consumer knowledge of the energy content of alcoholic drinks. See Table 1. Studies were conducted across a range of continents (Europe, US and Australasia). All studies were rated as low quality. Knowledge of the energy content of wine was examined in 3 studies and the remaining 5 studies examined knowledge of energy content of a range of alcoholic drinks. Seven studies reported results for knowledge of energy content independent to other nutrients and a single study reported knowledge for nutritional content (including calorie content). Studies typically asked participants to estimate the number of calories in drinks using a self-report questionnaire format (e.g. ‘How many calories are in a glass of red wine?’). Across studies it was common for a sizeable proportion of participants to be inaccurate in their estimation of calorie content (38-83% of participants across studies). A sub-set of studies reported on the direction of inaccuracy (n=6). In 4/6 studies it was most common for participants to overestimate energy content (i.e. believe there were more calories in drinks than in reality) and in 2/6 studies underestimation was reported to be more common. One high quality experimental study that was designed to examine the impact of energy labels on consumer behaviour ^29^ included estimation of calorie content as a secondary outcome and reported data on direction of calorie estimation inaccuracy in a group of participants not exposed to energy labels; participants tended to overestimate calorie content (see Table 3). We were able to meta-analyse nine effects from six studies (see Figure 2 and online supplementary materials for more detail). The pooled proportion of participants accurately estimating energy content was 26% (95% CI: 18% - 36%), with considerably high levels of heterogeneity (I^2^ = 97%). Leave-one-out analyses demonstrated that results were not markedly affected by the inclusion of any one study (smallest = 24%, largest = 28%). We also examined and adjusted analyses for potential publication bias and results remained unchanged. See online supplementary materials. Based on the consistency of findings and directness of evidence, but relatively small number of mainly low-quality studies, we concluded that there is moderate evidence that people tend to be unaware of the energy content of alcoholic drinks.

**Table 1.** Studies examining knowledge of energy content of alcoholic drinks.

| Study | Sampling approach | Participant information | Main review question(s) addressed | Main outcome variable(s) | Experimental manipulation | Results | Overall quality rating |
| --- | --- | --- | --- | --- | --- | --- | --- |
| Annunziata, 2016 (a)<sup>25</sup> | 300 Italian wine consumers<br>Online and in-person surveys | 51.5%F, 48.5%M<br>Aged 18 +<br>No BMI data<br>48% University educated<br>Monthly wine drinkers | Knowledge of energy content of alcoholic drinks | Self- report questionnaire<br>'How many kcal are contained in a glass of red Wine?' | N/A | 20% of sample identified correct kcal content<br>60% of sample underestimated kcal content.<br>20% of sample overestimated kcal content | Low |
| Annunziata 2016 (b)<sup>26</sup> | 1016 wine consumers from Italy, France, Spain and the USA (east coast)<br>Online survey | 51%F, 49%M<br>Aged 18 +<br>No BMI data<br>38% University educated<br>Monthly wine drinkers | Knowledge of energy content of alcoholic drinks | Self- report questionnaire<br>'How many kcal are contained in a glass of red Wine?' | N/A | Italy: 22% of sample accurate<br>63% of sample underestimated<br>Spain: ~30% of sample accurate.<br>50% of sample underestimated<br>France: 36% of sample accurate<br>Direction of inaccuracy not reported<br>US: 28% of sample accurate<br>43% of sample underestimated | Low |
|  |  |  |  |  |  | 29% of sample overestimated |  |
| Bui, 2008 (Pilot Study)<sup>8</sup> | 58 US undergraduate students<br>Data collection method not reported | 58%F, 42%M<br>Aged 20-33<br>No BMI info<br>All University educated<br>85% had drank alcohol in the past month | Knowledge of energy content of alcoholic drinks | Self-report measure<br>Participants estimated the absolute levels of calories for standard drink sizes of beer (12oz), wine (5oz) and distilled liquor (1.5oz)<br>Wording of question not reported | N/A | For all beverages, percent accuracy = 21%, participants overestimated the energy content, but no direct statistical comparison reported | Low |
| Grunert, 2018<sup>36</sup> | 5,395 participants from Denmark, Germany, UK, Spain, Netherlands and Poland.<br>Online survey | ~50%F, 50%M<br>Aged 18-65<br>No BMI info<br>12-41% University educated<br>No information on frequency of drinking any alcohol | Knowledge of energy content of alcoholic drinks | Self-report measure<br>'Product knowledge about nutrition was measured by asking respondents about the content of calories, fat and carbohydrates of alcohol-free beer, regular beer (between 4.5 and 5.5% alcohol), white wine, red wine, and whiskey. Answering options for calories were intervals of 50 kcals up to >300 kcals | N/A | Nutrition knowledge tended to be low, with participants on average incorrectly estimating nutritional content of drink types | Low |
| Pabst, 2019<sup>32</sup> | 21 German wine consumers<br>Focus group | 48% F, 52% M<br>Aged 18+<br>No BMI info<br>76% University educated<br>All consumed wine twice monthly or more | Knowledge of energy content of alcoholic drinks | Self-report measure<br>Participants were asked to estimate the amount of calories in a sample of wine and to write down their estimate for wine and other alcoholic beverages | N/A | 76% of the answers were wrong. Most participants (16/21) overestimated energy content | Low |
| Vecchio, 2018<sup>34</sup> | 103 Italian wine consumers<br>Laboratory study | 51% F, 49% M<br>M age = 29.1 (SD 7.1)<br>No BMI info<br>60% university educated<br>All participants consumed wine once a week or more | Knowledge of energy content of alcoholic drinks | Self-report measure<br>Participants were asked to indicate the kcal content of different types of wine (white and sweet wine) through a multiple-choice question. Exact question wording not reported. | N/A | For white wine, 38% were incorrect in estimating kcal content, 62% were correct<br>For sweet wine, 69% were incorrect in estimating kcal content, 31% were correct | Low |
| GfK<br>(Consumer research company),<br>2014<sup>35</sup> | 5395 European participants<br>(Germany, Poland, Denmark, Netherlands, Spain, UK)<br>Online survey | 51%F, 49%M<br>Aged 18 +<br>No BMI data<br><85% University educated<br>No info on alcohol drinking habits | Knowledge of energy content of alcoholic drinks | Self-report questionnaire<br>“How many calories (in kcal) do you think are provided by each of the following products?” Beer, white wine, red wine, whiskey, with response options in 50kcal intervals from <50 to >300 | N/A | For all drink types 30-33% participants chose answer ‘I don’t know’.<br>For all drink types only 6-17% of participants selected correct option.<br>For all drink types, overestimation of calorie content was most common (50-96%) participant response | Low |
| Alcohol Research UK Report<br>(Maynard), 2018<sup>38</sup><br>Study 1 | 450 UK alcohol drinkers<br>Online study | 54%F, 46%M<br>Aged 18+ (M = 38)<br>No BMI info<br>65% University educated<br>All had previously consumed alcohol | Knowledge of energy content of alcoholic drinks | Self-report measure<br>Participants were asked to estimate the number of calories and units in a selection of alcoholic drinks (cider, beer, alcopops, wine, gin & tonic) | N/A | Calories in alcoholic drinks were consistently over-estimated | Low |

**Figure 2.**
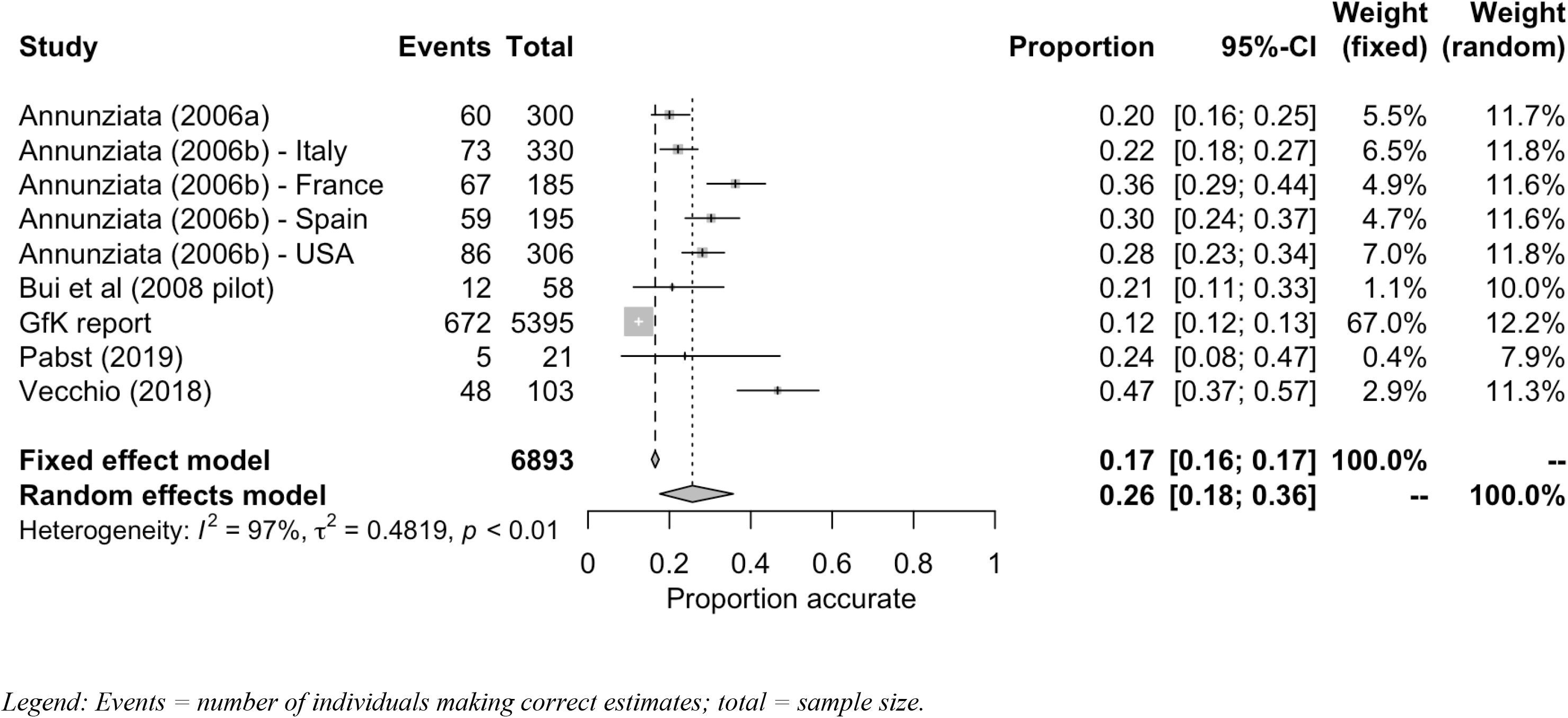
Meta-analysis of studies examining proportion of participants accurately estimating energy content of alcoholic beverages.

### Support for energy labelling of alcoholic drinks

Nine studies examined consumer support for energy labelling of alcoholic drinks (see Table 2). Studies were conducted across a range of continents (Europe, US and Australasia). Study quality tended to be low (n=6), with a minority of studies rated as moderate quality (n=3). Support for energy labelling of wine was examined in two studies and the remaining seven studies examined support for labelling of alcoholic drinks in general. Three studies reported on support for energy information labelling only and six studies reported on support for nutrition information (including calories) labelling. Studies typically measured support for energy labelling using a self-report questionnaire format (e.g. “it should be a requirement that nutritional information is displayed on bottles/cans/casks of alcohol”). It was common for a sizeable proportion of participants to support labelling of alcoholic drinks (41-84% of participants across studies). Four studies reported data on the number of participants supporting vs. opposing labelling. In all of these studies it was more common for participants to support rather than oppose labelling. One experimental study, which was rated as moderate quality, that examined the impact of energy labels on consumer behaviour ^39^ also included support for energy labelling as a secondary outcome and reported data on support for labelling in a group of participants not exposed to energy labels; the majority of participants supported labelling (see Table 3). We were able to meta-analyse ten effects from nine studies (See Figure 3 and online supplementary materials). The pooled proportion of participants supporting energy labelling was 64% (95% CI: 53% - 73%), with high heterogeneity (I^2^ = 99%). Leave-one-out analysis demonstrated that results were not markedly affected by the inclusion of any one study (smallest = 60%; largest = 67%). We also examined and adjusted analyses for potential publication bias and results remained unchanged. See online supplementary materials. Based on the consistency of findings and directness of evidence, but relatively small number of mainly low-quality studies, we concluded that there is moderate evidence that people are more likely to support than oppose energy labelling of alcoholic drinks.

**Table 2.** Studies examining support for energy labelling of alcoholic drinks.

| Study | Sampling approach | Participant information | Main review question(s) addressed | Main outcome variable(s) | Experimental manipulation | Results | Overall quality rating |
| --- | --- | --- | --- | --- | --- | --- | --- |
| Annunziata, 2016 (a)<sup>25</sup> | 300 Italian wine consumers<br>Online and in-person surveys | 51.5%F, 48.5%M<br>Aged 18 +<br>No BMI data<br>48% University educated<br>Monthly wine drinkers | Support for energy labelling of alcoholic drinks | Self-report questionnaire<br>Support for more nutrition info on wine labels.<br>Wording of question not reported | N/A | 55% reported it would be particularly useful to receive more information on nutritional and health features on wine labels | Low |
| Annunziata 2016 (b)<sup>26</sup> | 1016 wine consumers from Italy, France, Spain and the USA (east coast)<br>Online survey | 51%F, 49%M<br>Aged 18 +<br>No BMI data<br>38% University educated<br>Monthly wine drinkers | Support for energy labelling of alcoholic drinks | Self-report questionnaire<br>'How interested are you in receiving nutrition information on wine labels?' 1 (not at all) to 5 (totally) scale | N/A | Italy: M=3.4 (SD 1.2), indicating of some interest<br>Spain: M=2.9 (SD 1.2), indicative of some interest<br>France: M=2.2 (SD 1.2), indicative of some interest<br>US: M=3.6 (SD 1.3), indicative of some interest | Low |
| Christensen, 2019<sup>27</sup> | 6,000 Danish participants<br>Online survey | 50%F, 50%M<br>Aged 18-74<br>No BMI info<br>47% higher education level<br>64% consume | Support for energy labelling of alcoholic drinks | Self-report measure<br>Support for mandatory nutrition labeling | N/A | 46% of participants reported that mandatory nutrition labelling was a | Moderate |
|  |  | alcohol during a typical week |  | on alcohol.<br>Response options: “very good suggestion”, “good suggestion” “neither good or bad suggestion”, “bad suggestion”, “very bad suggestion”, “I do not know” |  | good suggestion or very good suggestion |  |
| Kypri, 2007<sup>28</sup> | 7224 Australian students<br>Online survey | No gender info<br>Aged 17-25<br>No BMI info<br>All in higher education<br>90% consumed alcohol in the past year | Support for energy labelling of alcoholic drinks | Self-report measure<br>‘It should be a requirement that nutritional information (eg, the amount of sugar and kilojoules) is displayed on bottles/cans/casks of alcohol’<br>Strongly agree to Strongly disagree response options | N/A | 40% strongly agreed<br>35% agreed<br>20% neither agreed or disagreed<br>3% disagreed<br>1% strongly disagreed | Moderate |
| Moore, 2010 (Shape Up American! Survey)<sup>30</sup> | 503 adult Americans<br>Online survey | No gender info<br>Aged 18 +<br>No BMI info<br>No education level info<br>No information on frequency of drinking any alcohol | Support for energy labelling of alcoholic drinks | Self-report<br>‘How Important Is It to You to Have Each of the Following Types of Information on an Alcoholic Beverage Label?’ | N/A | 84% of participants reported that including calorie information was either important or somewhat important | Low |
|  |  |  |  | The number of calories in each drink. Response options not fully reported |  |  |  |
| Nikolaou, 2014<sup>31</sup> | 1440 Scottish Undergraduate students<br>Online survey | 67%F, 33%M<br>M age = 20.3 (SD 2.9)<br>M BMI = 23.0 (SD 4.6)<br>All in higher education<br>No information on frequency of drinking any alcohol | Support for energy labelling of alcoholic drinks | Self-report measure<br>Multiple choice question on calorie labelling on alcohol. Wording of question not reported | N/A | Half of female participants and a third of males reported they would like to see calorie information on alcohol | Low |
| Thomson, 2012<sup>33</sup> | 1523 Australian adults<br>Telephone interview | No gender info<br>Aged 16 +<br>No BMI info<br>No education info<br>No info on frequency of drinking any alcohol | Support for energy labelling of alcoholic drinks | Self-report measure<br>Participants were asked to provide level of support for including nutritional information (energy, protein, fat, carbs, sugars) on alcoholic drink labels | N/A | 76% strongly support or support. 7% neither support nor oppose. 17% strongly oppose or oppose | Moderate |
| GFK (Consumer research company), 2014<sup>35</sup> | 5395 European participants (Germany, Poland, Denmark, Netherlands, Spain, UK)<br>Online survey | 51%F, 49%M<br>Aged 18 +<br>No BMI data<br><85% University educated<br>No info on alcohol drinking habits | Support for energy labelling of alcoholic drinks | Self-report questionnaire<br>Level of agreement with "The same nutrition information (energy value, proteins, carbohydrates, sugars, fat, saturated fats, salt) should be provided for all food and drink products (alcoholic and non-alcoholic)", with response format strongly disagree (1) to strongly agree (7) | N/A | 69% of participants agreed with statement, 16% were neutral, 14% disagreed | Low |
| Alcohol Research UK Report (Maynard), 2018<sup>38</sup><br>Study 1 | 450 UK alcohol drinkers<br>Online study | 54%F, 46%M<br>Aged 18 + (M = 38)<br>No BMI info<br>65% University educated<br>All had previously consumed alcohol | Support for energy labelling of alcoholic drinks | Self-report measure<br>Participants reported whether they agreed or disagreed with the statement "Calorie information on alcoholic drinks is a good idea" | N/A | 81% of participants agreed that calorie information on alcohol drinks is a good idea. 7% disagreed | Low |

**Figure 3.**
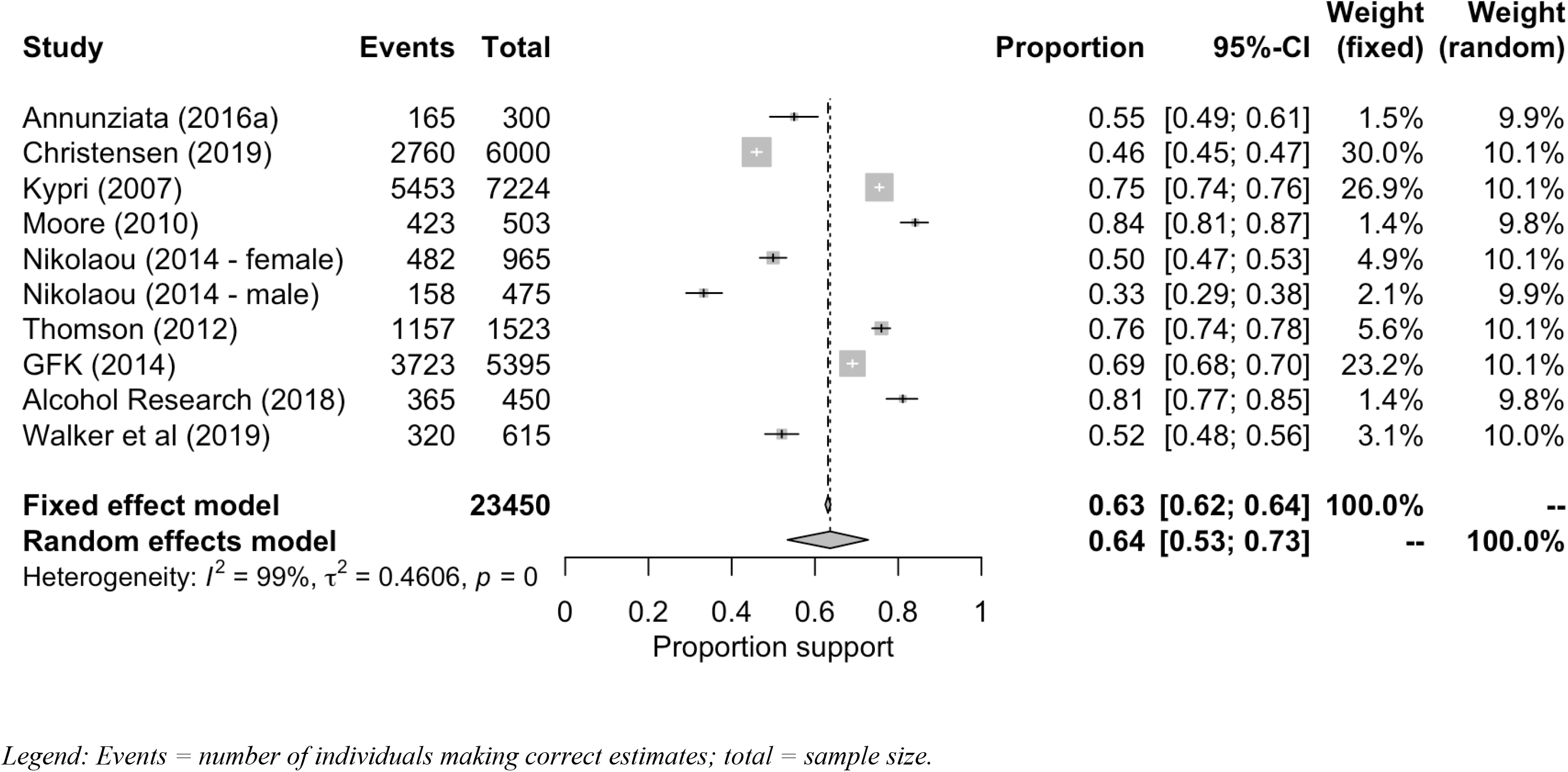
Meta-analysis of studies examining proportion of participants supporting energy labelling policy.

### Effects of energy labelling on consumer behaviour

Six studies examined the effects of energy labelling on consumer-related behaviour (see Table 3) and as differing methodologies were adopted, we summarise studies individually. Maynard et al. ^29^ examined the effect of providing information about the calorie content of beer vs. no calorie content information in a group of UK participants recruited from a University setting. The main outcome measure used was the volume consumed from a glass of beer that was served during a mock taste-test in a laboratory setting. A secondary outcome measure was intended future consumption of the beer. There was no effect of labelling on beer consumption in the taste-test or on intended future consumption. We deemed the quality of the study to be high. However, the outcome measure did not involve participants choosing a drink or making decisions about how many drinks to order/consume and therefore the study was unable to measure a number of pathways by which drinking behaviour may be affected by energy labelling.

**Table 3.**
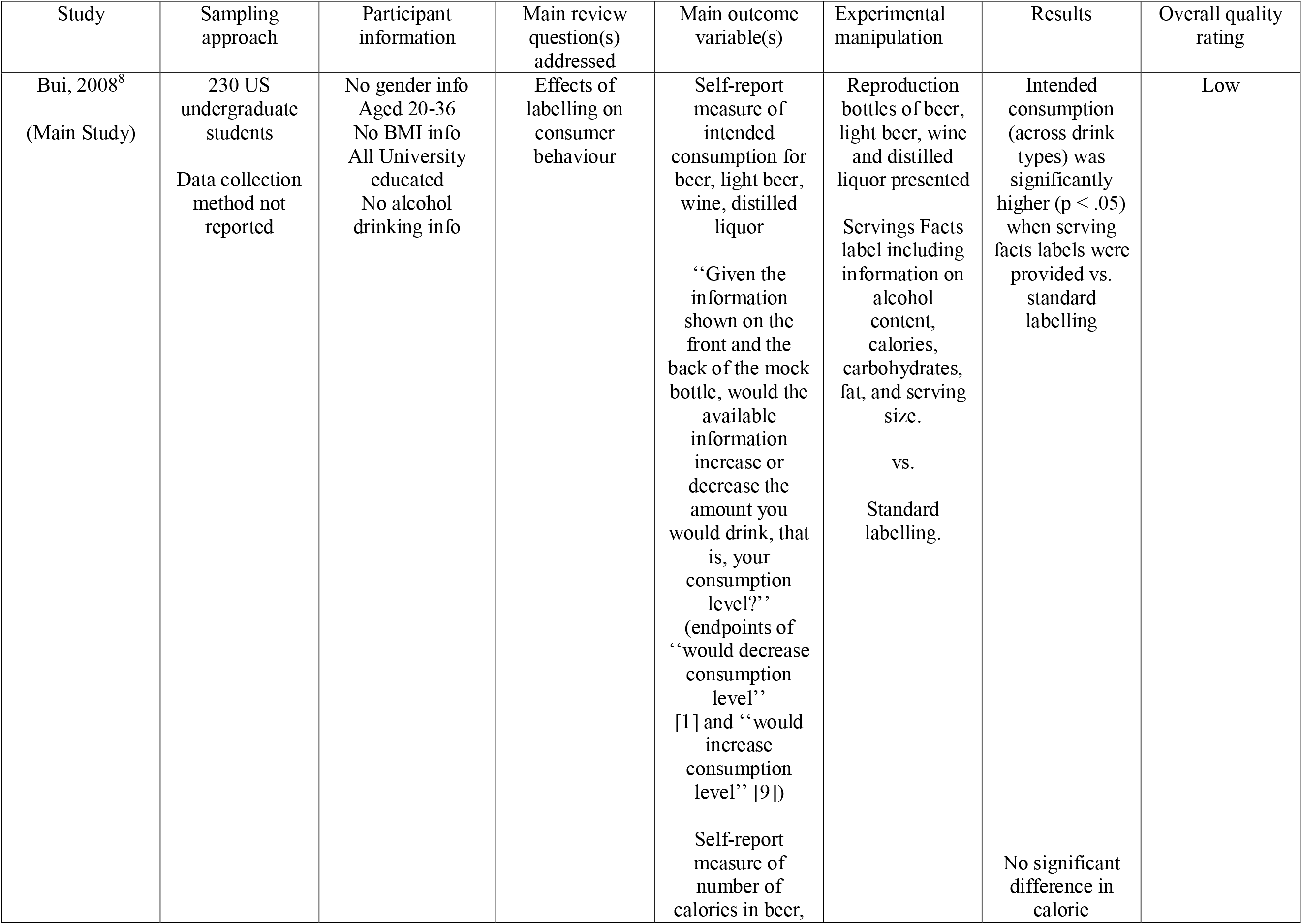
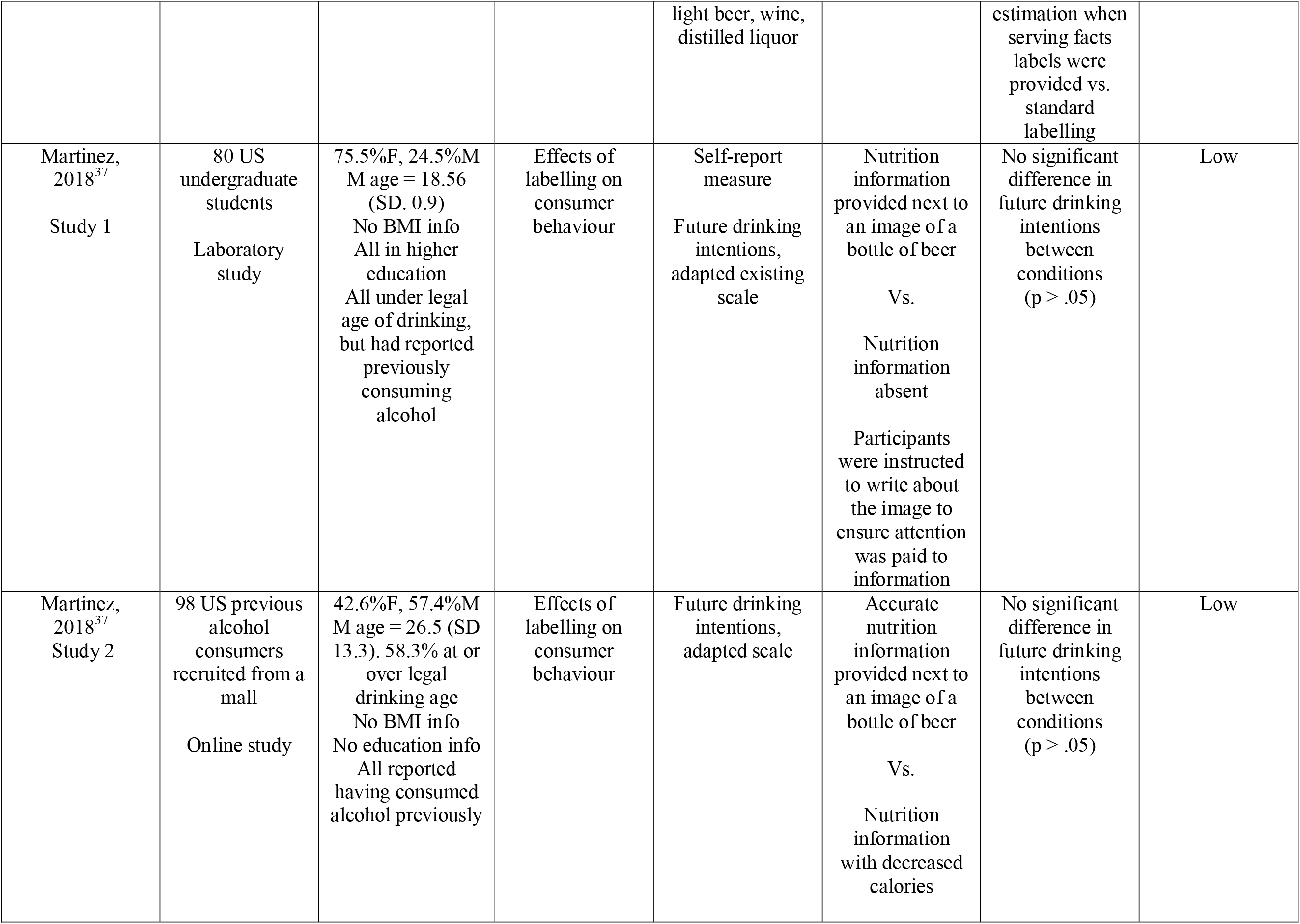
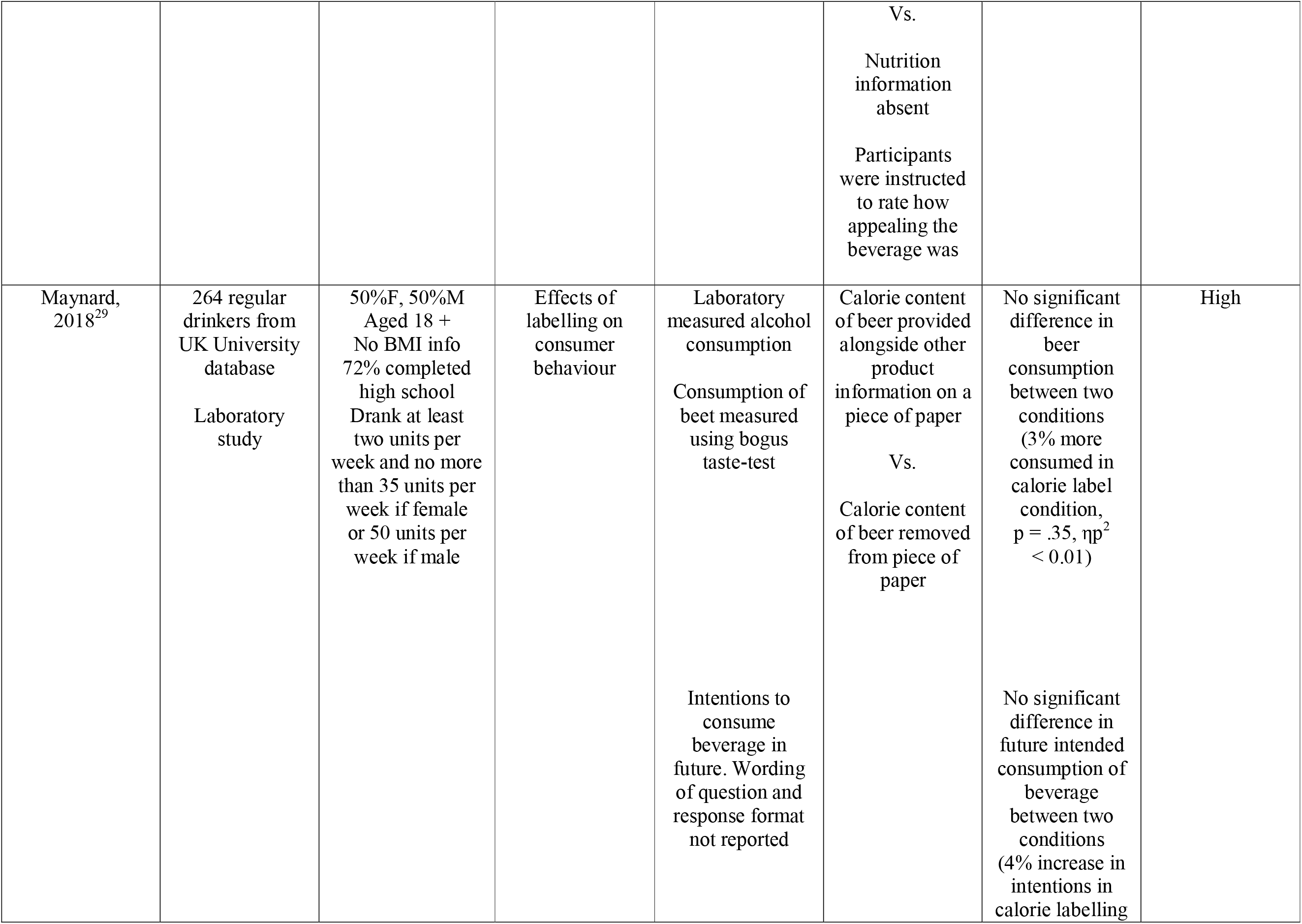
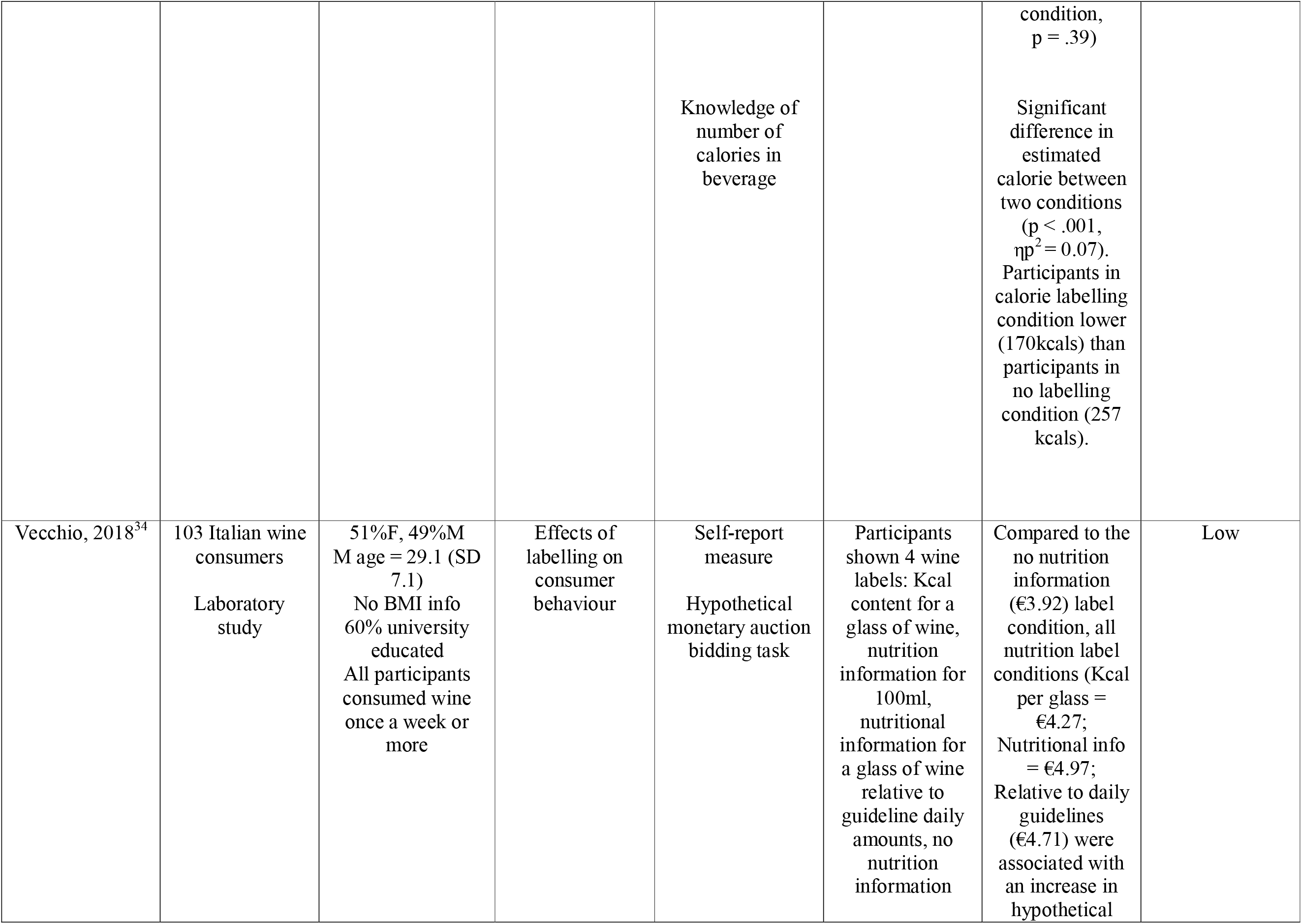
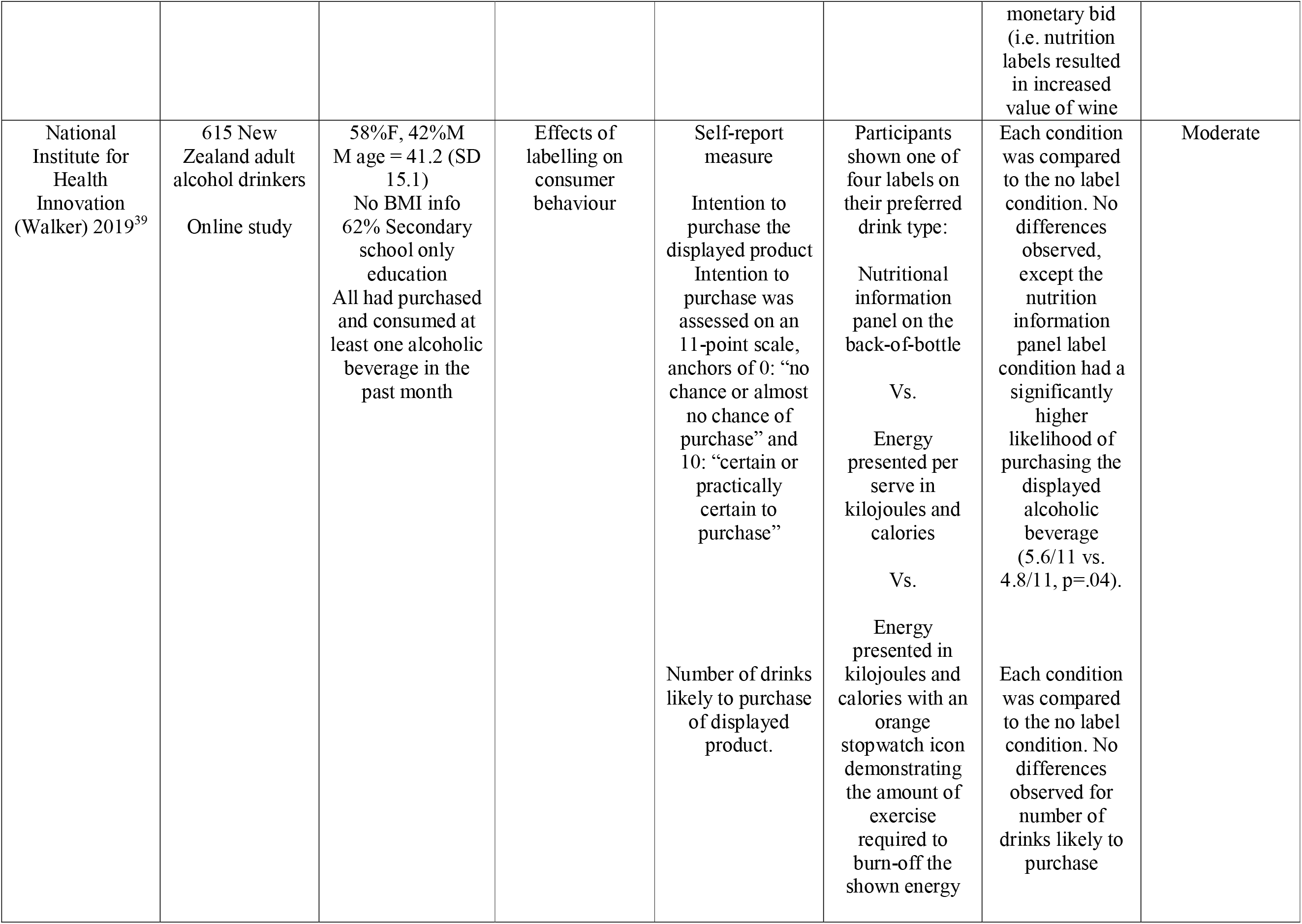
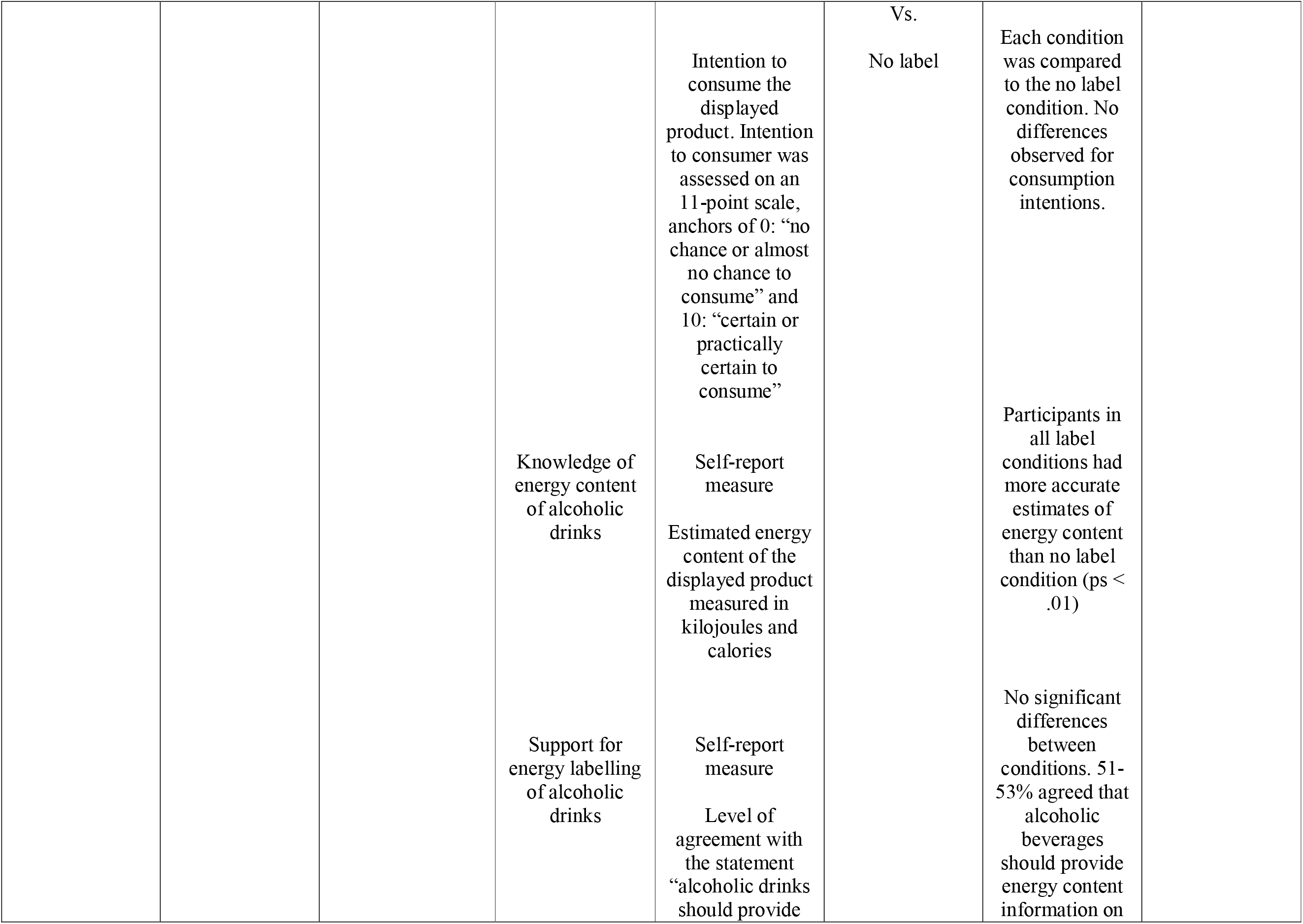
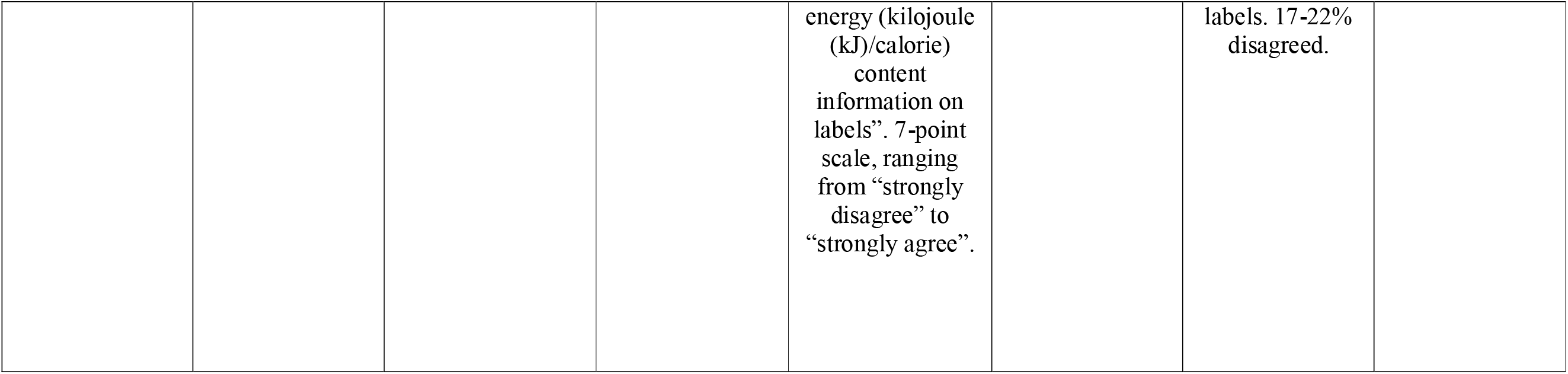
Studies examining effects of labelling on consumer behaviour.

In a moderate quality study of participants from New Zealand, Walker et al. ^39^ examined the effect that nutrition labelling conditions (nutrition information panel vs. calories and kilojoules per serving vs. calorie and kilojoules per serving plus exercise required to burn off energy vs. no nutrition information) had on a range of self-reported intention measures (intention to purchase, number of drinks likely to purchase, intention to consume). There were no significant differences between any of the nutrition information conditions vs. no nutrition information for intended consumption or number of drinks likely to purchase. For intended purchasing, the nutrition information panel condition had a significantly higher likelihood of purchasing the displayed alcoholic beverage relative to the no information condition. The calorie and kilojoule information conditions (with or without exercise information) did not significantly differ to the no nutrition information condition.

Vecchio et al. ^34^ sampled Italian wine consumers and examined the amount of money participants were willing to bid (hypothetical) in a mock auction bidding task for wine products that were labelled with calorie content per glass, full nutrition information (per 100ml or per glass), guideline daily amount labelling (with key nutrients), or with no nutrition information. Participants were exposed to all label conditions (repeated-measures study). Compared to the no nutrition information, all labelling conditions (including the calorie content label condition) were associated with a significantly higher hypothetical monetary bid. Study quality was rated as low. We deemed that the outcome measure was eligible for inclusion as it may act as a proxy measure of intended purchasing behaviour. However, given the hypothetical nature of the measure and that participants were exposed to all conditions, the findings may reflect a tendency to report that any additional product information (as opposed to limited product information) increases perceived monetary value, as opposed to nutrition information increasing purchasing intentions for wine.

In a low-quality study of US University students, Bui et al. ^8^ examined the effect that the inclusion of calorie information labelling (alongside other nutrition information) vs. no nutrition information labelling on a range of alcoholic drinks had on self-reported intended behaviour (‘Would the information increase or decrease the amount you would drink?’). Across all drink types, intended consumption did not differ in the labelling vs. no labelling condition.

Martinez ^37^ conducted two studies; examining the effect that nutrition information on a bottle of beer (vs. no nutrition information) had on self-reported future drinking intentions. The first study was of US university students and the second was of US adults recruited in a shopping mall. Both studies were rated as being low in quality and there was no significant effect of nutrition information on future drinking intentions in either study.

The majority of studies (including the one rated high-quality study) found evidence to suggest that energy labelling did not have an effect on consumer behaviour, via actual (consumption) or proxy measures (intentions) of consumer behaviour. There were some inconsistencies in findings and studies tended to use self-reported hypothetical measures (lack of directness of evidence) of alcohol consumption. As such, the overall quality of evidence supporting this conclusion was rated as very low.

### Other outcomes

Three of the experimental studies on consumer behaviour also examined whether calorie estimates for alcoholic drinks were affected by exposure to labelling (vs. no labelling). In Bui et al. (low quality) there was no effect on calorie estimation. In Maynard et al. (high quality) provision of calorie labels had a significant effect on calorie estimation and directionally calorie estimates improved (relative to a no calorie label condition). In Walker et al. (moderate quality) participants in all nutrition label conditions had significantly more accurate estimates of calorie content than those in the no label condition. Walker et al. also examined support for energy labelling and found that this was not affected by exposure to labelling.

## Discussion

In the present research we conducted a rapid systematic review to assess evidence from studies examining consumer knowledge of the energy content of alcoholic drinks, support for energy labelling of alcoholic drinks and experiments examining the effect of energy labelling of alcoholic drinks on consumption behaviour.

Eight studies examined consumer knowledge of the energy content of alcoholic drinks. Although study quality tended to be low, findings were consistent across studies and a substantial proportion of participants in all studies were inaccurate when asked to estimate the number of calories in different alcoholic drinks. In a meta-analysis of nine effects from six studies addressing this question, the pooled proportion of participants accurately estimating energy content of alcoholic drinks was 26% (95% CIs 18% - 36%). Based on these findings and the relatively small numbers studies addressing this question we graded the overall level of evidence in support of this conclusion as ‘moderate’. However, the extent to which inaccuracy when estimating calorie content was more likely to be caused by overestimation (more calories perceived) or underestimation (fewer calories perceived) was not consistent across studies. This consideration may be important because it has been suggested that if consumers expect a product to have higher calories than is presented on nutrition labelling this may cause a ‘backfiring’ effect of increased consumption ^40^. Given the relatively small number of studies addressing this question, future research identifying factors relating to the direction and size of the misperception of alcoholic drink energy content would therefore be informative. However, it should also be noted that directionality of misperception may not be the most important factor affecting the extent to which calorie information impacts on consumer behaviour. The presence of calorie information may also serve to remind or ‘prime’ consumers about the importance of limiting their energy intake ^41^ or allow consumers to choose relatively lower calorie drink options, neither of which rely on the assumption that consumers only change their behaviour in response to labelling because they tend to under/overestimate calorie content.

Nine studies examined consumer support for energy labelling of alcoholic drinks. Studies tended to be of low quality, but it was consistently found that a sizeable proportion of sampled participants supported calorie and nutritional information being provided on labelling of alcoholic drinks. Of the smaller number of studies that reported the numbers of participants supporting vs. opposing labelling, it was more common for people to support labelling than oppose it. Studies included in the present review that contributed evidence to consumer support for energy labelling were from a range of countries (US, European countries, Australia, New Zealand) and across these studies there was consistent support for energy labelling. In a meta-analysis of ten effects from nine studies, the pooled proportion of participants supporting energy labelling was 64% (95% CI: 53% - 73%). Based on these consistent findings, but the relatively small number of studies we graded, the overall level of evidence in support of this conclusion as ‘moderate’. These findings are in line with other research which has found that the general public are likely to support public health policies that involve information provision in order to improve health ^42 43^.

Six studies examined the effects of energy labelling of alcoholic drinks on consumer behaviour related outcomes. Studies tended to be of low quality and relied on self-reported proxy measures of alcohol consumption (e.g. intended consumption).Although intended alcohol consumption tends to correlate with actual consumption ^44^, it is well recognised that intentions will often not be followed by successful behaviour change ^45^ and the extent to which intentions do predict alcohol drinking are likely moderated by individual difference and contextual factors ^46^. Therefore, the reliance in included studies to reply on proxy measures such as intentions is a significant limitation. One high quality study examined actual consumption of alcohol, although this was conducted in a laboratory setting and the outcome measured was amount of a served beverage consumed in a mock taste-test ^47^. Energy labelling may impact consumer behaviour by altering drink choice or reducing the number of drinks ordered, but no studies examined this. In addition, energy labelling of alcoholic drinks may impact on consumer behaviour by affecting diet (e.g. eating less during, before or after drinking) or increase energy expenditure by increasing physical activity ^48^.

Given these considerations and the small number of largely low-quality studies conducted, we concluded that at present there is very low-quality evidence which suggests that energy labelling of alcoholic drinks does not affect consumer behaviour. However, this conclusion could be changed by contradictory findings and there a need for higher quality studies of the impact that energy labelling of alcoholic drinks has on behaviour (e.g. alcohol consumption and overall energy intake vs. expenditure). Understanding whether there are any unintended consequences of energy labelling of alcoholic drinks (e.g. meal skipping resulting in increased harm from drinking) will also be important. Although some of the studies included in the present review reported information relating to socioeconomic position (SEP) of sampled participants, outcomes were not routinely reported based on SEP. Because there is some evidence that information provision policies, like nutrition labelling, may exert a stronger influence on the behaviour of people of higher SEP and therefore create inequality ^49 50^, future research on the effects of energy labelling of alcoholic drinks would benefit from considering equity of intervention effectiveness.

We conducted a rapid rather than a standard systematic review in order to be able to inform a policy consultation on the mandatory labelling of alcoholic products in the UK ^10^. We adopted similar methodology to other rapid evidence reviews and followed best practice guidelines ^17 18^, but there are limitations to this approach. We searched two suitable electronic databases and it is plausible that we may have found more studies if we had searched more. To mitigate this, we conducted forward citation tracking, searched the reference list of all eligible articles and included studies reported by public health bodies but not published in journals. We retained a number of important methodological features of standard systematic reviews, including independent full-text screening of articles for eligibility by two researchers and coding of study quality by two researchers independently. We examined evidence of publication bias in the limited studies we were able to quantitatively synthesise, but given that we were unable to meta-analyse all study outcomes and we were only able to include a small number of studies which limits statistical power of formal tests of publication bias. Due to the relatively small number of eligible studies, unpublished high-quality studies may change conclusions made.

There were a number of limitations to the studies included in the review. Study quality tended to be low and this was often because of incomplete reporting of study methodology and results. Most studies did not report information on potential conflicts of interest and pre-registration of methodology and analysis plans was reported in only 1/16 studies. Four of the included studies were not published in peer-reviewed journals and instead were published and made available online by public health bodies and research institutes. However, findings from these studies were consistent with those published in journals. The present research focused on consumer behaviour, but any evaluation of the effectiveness of energy labelling of alcoholic drinks as a public health measure will require a global approach that also examines industry behaviour. For example, there is some evidence that the provision of nutrition information about food products may result in food manufacturers reducing the energy content of food products ^51^. A similar process could occur with alcoholic drink manufacturers and reductions to energy content could be achieved through introducing new products ^52^, or reducing existing serving size and/or alcohol content by volume (ABV). As both reductions to alcoholic beverage serving sizes ^53^ and ABV^54^ decrease alcohol consumption, industry reformulation as a result of energy labelling may be beneficial to public health.

### Conclusions

There is a moderate level of evidence that people tend to be unaware of alcoholic drink energy content and are more likely to support than oppose energy labelling of alcoholic drinks. Currently evidence suggests that energy labelling of alcoholic drinks is unlikely to directly affect consumer behaviour. However, this conclusion is based on a very small number of studies with substantial methodological issues (very low evidential value) and may change as a result of higher quality studies conducted in real-world settings.

## Data Availability

The pre-registered protocol for the review is available at https://osf.io/8gpm5/ and data tables are reported in the appendix.

